# Analysis of attitude, behavior intention and influencing factors of community medical staff on respite care

**DOI:** 10.64898/2026.01.06.26343503

**Authors:** Qi-Ni Pan, Li-Chong Lai, Hui-Qiao Huang, Hong-Ju Deng, Ling-Li Liang, Cai-Li Li, Deng-Jing Fan, Dong-Na Zhou, Yi-Fen Liao

**Affiliations:** Emergency department, The Second Affiliated Hospital of Guangxi Medical University, Guangxi, China; Party Committee Office, The Second Affliated Hospital of Guangxi Medical University, Guangxi, China; General Surgery, The Second Affliated Hospital of Guangxi Medical University, Guangxi, China; Nursing Department, The Second Affliated Hospital of Guangxi Medical University, Guangxi, China

**Author notes:** Correspondence: Dong-Na Zhou, Yi-Fen Liao;, Adress: No. 166 Daxue East Road, Xixiangtang District, Nanning City, Guangxi, China; 530007. co-first author: Qi-Ni Pan, Li-Chong Lai.

**Keywords:** community medical staff, respite services, attitude, behavioral intention, influencing factors

## Abstract

**Objective:** To investigate the attitude and behavior intention of community medical staff towards respite services and analyze its influencing factors, so as to provide reference for the development of respite services.

**Methods:** A multi-stage stratified cluster sampling method was used to select 348 community medical staff to conduct a survey through the questionnaire star. Single factor and multiple linear regression were used to analyze the factors affecting the attitude and behavior intention of community medical staff.

**Results:** The attitude score of community medical staff was 43.64, the behavioral intention score was 27.79, and the attitude and behavioral intention scores were low. The results of multivariate linear regression analysis showed that educational background, occupation, relationship with the elderly at home, whether there were family members who could not take care of themselves due to major diseases, whether they had the experience of taking care of the elderly, and whether they had received knowledge training in respite services were the factors affecting the attitude of community medical staff to respite services ( P < 0.05 ). Educational background, working years, whether there are family members who cannot take care of themselves due to major diseases, whether they have the experience of taking care of the elderly, whether they know the respite services, whether they have received knowledge training in respite services and whether they know the respite services are the factors affecting the behavioral intention of community medical staff (P < 0.05).

**Conclusion:** The attitude of community medical staff towards respite services is not positive, and their behavioral intention is low. There are many factors affecting attitude and behavioral intention. It is necessary to strengthen the training of respite services knowledge for medical staff with low education and short working years, strengthen the cultivation of empathy and empathy ability of medical staff, carry out multi-level publicity of respite services, and improve people’s awareness and acceptance of respite services, so as to promote the development of respite services industry and promote healthy aging.

## 1. Introduction

In many countries and regions, the proportion of the elderly population continues to rise. By 2030, the global population aged 60 and above will reach 1.4 billion, and by 2050, it will reach 2.1 billion.[1]∘Due to the gradual decline of physical function, the elderly population often becomes the high-risk group and the main group of people with various chronic diseases or disabilities.[2]∘The results of the fourth sample survey on the living conditions of the elderly in urban and rural areas in China show that there are 150 million elderly people with chronic diseases, accounting for 65 % of the total number of the elderly, including more than 40 million disabled elderly people with limb dysfunction and cognitive decline[3]. Chronic diseases such as hypertension, diabetes, coronary heart disease, and malignant tumors have a high incidence in the elderly, and these diseases usually require long-term treatment and care, making the elderly face many difficulties in daily life[4, 5]. In this case, family members assume the main responsibility for care, but long-term care not only consumes a lot of time, energy and economic costs, but also may lead to the physical and mental health of caregivers themselves, and the burden of family care is increasingly heavy[6]. There is an urgent need for effective social support and professional service intervention to alleviate this dilemma.

Respite Care, also known as intermittent care service and temporary care service, refers to a service method that temporarily sends the elderly who are taken care of at home to a respite services institution or asks them to provide home care. Professional caregivers provide temporary, periodic and planned care services, so that their family caregivers can take a short break to prevent their physical and mental adverse reactions[7]. Studies have shown that respite services can improve the quality of life of the main caregivers of disabled elderly, alleviate the care burden of caregivers, and avoid health problems or psychological collapse due to long-term overworked care[8, 9]. Meanwhile, the respite service is provided by professionals, which can ensure that the elderly get scientific and reasonable care during the service period, meet their needs of life care, rehabilitation care, psychological comfort and other aspects, avoid unnecessary medical expenses caused by unprofessional care, and reduce the cost of family care to a certain extent[10, 11]. As a new pension service model, respite service has been proved to be an effective auxiliary method to deal with population aging by many studies at home and abroad.

However, the development of China’s respite service still faces obstacles. On the one hand, the public’s awareness and acceptance of respite services are relatively low, and they do not link their need for rest with the use of respite services. Some family caregivers have concerns about the care of the elderly to others, worrying about the quality of service and safety issues[12]. On the other hand, the supply system of respite services is not perfect, the number of professional caregivers is insufficient and unevenly distributed, and the service facilities and resources fail to meet the core needs of caregivers[13, 14]. In this case, grassroots medical institutions, as the basic link of the medical and health service system, are widely distributed, close to the community and residents, and can provide services for the elderly conveniently and quickly, with unique advantages and key roles[15]. At the same time, grassroots medical staff have professional medical care knowledge and skills, which can provide strong technical support for respite services. In multiple national policies, it is also emphasized that there is a need to support communities and institutions in providing family caregiver training and respite services for families of older adults with disabilities.

As the main body of providing respite services, the attitude and behavior intention of community medical staff directly affect the implementation and utilization of respite services[15]. In our study, the attitude and behavioral intention of community medical staff to respite service were investigated, and the influencing factors were analyzed, aiming to provide reference for the development of respite service. By understanding their attitudes and behavioral intentions, in-depth analysis of the current problems and obstacles that may exist in the process of carrying out respite services in primary medical institutions can provide scientific basis for formulating targeted policy measures and improve the enthusiasm of service participation, so as to promote primary medical institutions to better carry out respite services. It aims to improve the quality of life of the elderly and their families, improve the social pension service system, and promote the healthy development of the pension service industry.

## 2. Materials and Methods

### 2.1. Sample

From November 2024 to February 2025, a multi-stage stratified cluster sampling method was used. Firstly, according to the level of economic development and geographical location of Guangxi, Guangxi was divided into five layers : eastern Guangxi, northern Guangxi, western Guangxi, central Guangxi and Beibu Gulf coastal areas. Then, a questionnaire survey was conducted on the medical staff of community health service centers in 15 districts and counties in 3 urban districts and counties in each layer. Community medical staff inclusion criteria : age ≥ 18 years old ; working in the community for more than 1 year. Exclusion criteria : interns, trainees or those who do not want to participate in the survey. A questionnaire survey was conducted anonymously. After contacting the investigated unit by telephone and obtaining the consent of the corresponding person in charge, the content of the questionnaire was input into the questionnaire star website, and the two-dimensional code of the questionnaire was sent to the staff of the community health service center. Before filling in the questionnaire, they all need to read the questionnaire to fill in the requirements and guide them to answer the questions in a standardized way. The reading time is 1 min. After reading, they can be transferred to the home page of the questionnaire survey. Setting a mobile phone number can only fill in the questionnaire once, and all items can be submitted. A total of 372 questionnaires were distributed and recovered in our study, with 348 valid questionnaires. The study was approved by the Medical Ethics Committee of the hospital.

### 2.2. Data Measurement

Through literature review, expert consultation, combined with practical experience, a questionnaire was designed, including 3 parts, a total of 44 items. The first part is the general information questionnaire, including gender, age, nationality, educational background, working years, occupation, professional title, monthly income, physical condition, whether there are family members who cannot take care of themselves due to major diseases, whether they have the experience of taking care of the elderly, the relationship with the elderly at home, whether they have been engaged in the related work of the elderly health service in the past, whether they know the respite services, and whether they have received the knowledge training of the respite services. The second part is the attitude questionnaire of respite services, which mainly includes 14 items in 4 dimensions : the necessity of respite services, psychological activity, professional responsibility and respite services environment. Each item adopts the Likert 5-level scoring method, with ‘very agree, agree, neutral, disagree, strongly disagree’ accounting for 1 to 5 points respectively. The higher the score, the more positive the respite services attitude of the research object. The third part is the behavioral intention questionnaire of respite service, which mainly includes 10 items in 3 dimensions, such as active learning, providing service behavior and providing service content. The Likert 5-level score is used to ‘very unwilling to do, unwilling to do, uncertain, willing to do, very willing to do’. The higher the score, the better the behavioral intention of respite service.

### 2.3. Statistical Analysis

SPSS22.0 statistical software was used to analyze the data. The measurement data were expressed as ‘mean ± standard deviation’. The mean comparison between groups was performed by t test or analysis of variance. Multivariate analysis was performed by multiple linear regression model. P < 0.05 was considered statistically significant.

## 3. Results

### 3.1 Community medical staff respite services attitude, behavioral intention score

Our study included 348 community medical staff, 79 males and 269 females ; the mean age was 32.93 ± 8.20 years ( range, 19-50 years ). The working years ranged from 1 to 37 years, with an average of 10.13 ± 8.26 years. There were 99 doctors, 207 nurses and 42 others. The score of 348 community medical staff on the attitude of respite service was 22-57 points, with an average of 43.64 ± 5.64 points. The behavioral intention score was 11-48 points, with an average of 27.79 ± 4.93 points. Table 1.

**Table 1.** The scores of attitude and behavior intention of community medical staff ( n = 348 )

|  | entry | game |
| --- | --- | --- |
| attitude | The income and pay of respite service are not proportional to each other. | 2.31±0.64 |
| | Providing respite services makes me feel that my own value is not reflected. | 2.57±0.78 |
| | The respite service adds to our workload. | 2.76±0.47 |
| | respite services is too waste of time | 2.84±0.82 |
| | The respite service is not my job. | 2.88±0.93 |
| | Providing home respite services is not safe for our medical staff. | 2.90±0.68 |
| | respite services is of no significance | 3.02±0.85 |
| | Providing respite services for disabled older adults may be looked down upon. | 3.11±0.72 |
| | Providing respite services for disabled older adults can easily make me irritable and | 3.31±0.91 |
|  | depressed |  |
|  | Most elderly people who need respite services have poor home environment. | 3.38±0.84 |
|  | Respite services have no effect on the disabled elderly | 3.45±0.63 |
|  | When I came to provide respite services, it reflected the failure of medical staff. | 3.52±0.77 |
|  | The family atmosphere that needs respite service makes me feel uncomfortable. | 3.57±0.65 |
|  | The respite service does not solve the burden of primary caregivers. | 3.79±1.03 |
| behavioral<br>intention | I will assess the concerns and concerns of disabled elderly people and their families and provide targeted answers and counselling. | 3.02±0.63 |
|  | I will make every effort to provide respite services to the disabled elderly. | 3.07±0.99 |
|  | I will take the initiative to learn the relevant knowledge of respite services. | 3.06±0.76 |
|  | I will work hard to promote the development of respite service. | 3.08±0.56 |
|  | I will pay attention to the psychological feelings of the disabled elderly and their families and provide psychological care. | 3.14±0.83 |
|  | Pay close attention to families in need of respite services | 3.23±0.85 |
|  | I will provide rehabilitation guidance for the disabled elderly. | 3.41±0.77 |
|  | I will encourage my peers and volunteers to provide respite services for the disabled elderly. | 3.53±0.55 |
|  | I will take an active part in the practical training of respite service. | 3.55±0.91 |
|  | I will actively promote the benefits of respite services | 3.62±0.83 |

### 3.2 Single factor analysis of community medical staff’s respite services attitude and behavioral intention score

The results of single factor analysis showed that educational background, occupation, professional title, relationship with the elderly at home, physical condition, whether there were family members who could not take care of themselves due to major diseases, whether they had the experience of taking care of the elderly, and whether they had received knowledge training in respite services were the factors affecting the attitude of community medical staff to respite services ( P < 0.05 ). Occupation, educational background, working years, physical condition, family members with major diseases who could not take care of themselves, experience of taking care of the elderly, knowledge of respite services, knowledge training of respite services and knowledge of respite services were the factors affecting the behavioral intention of community medical staff ( P < 0.05 ). Table 2.

**Table 2.**
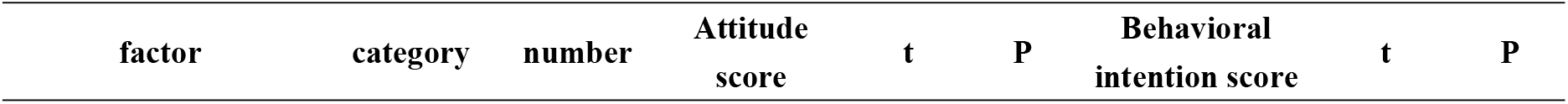
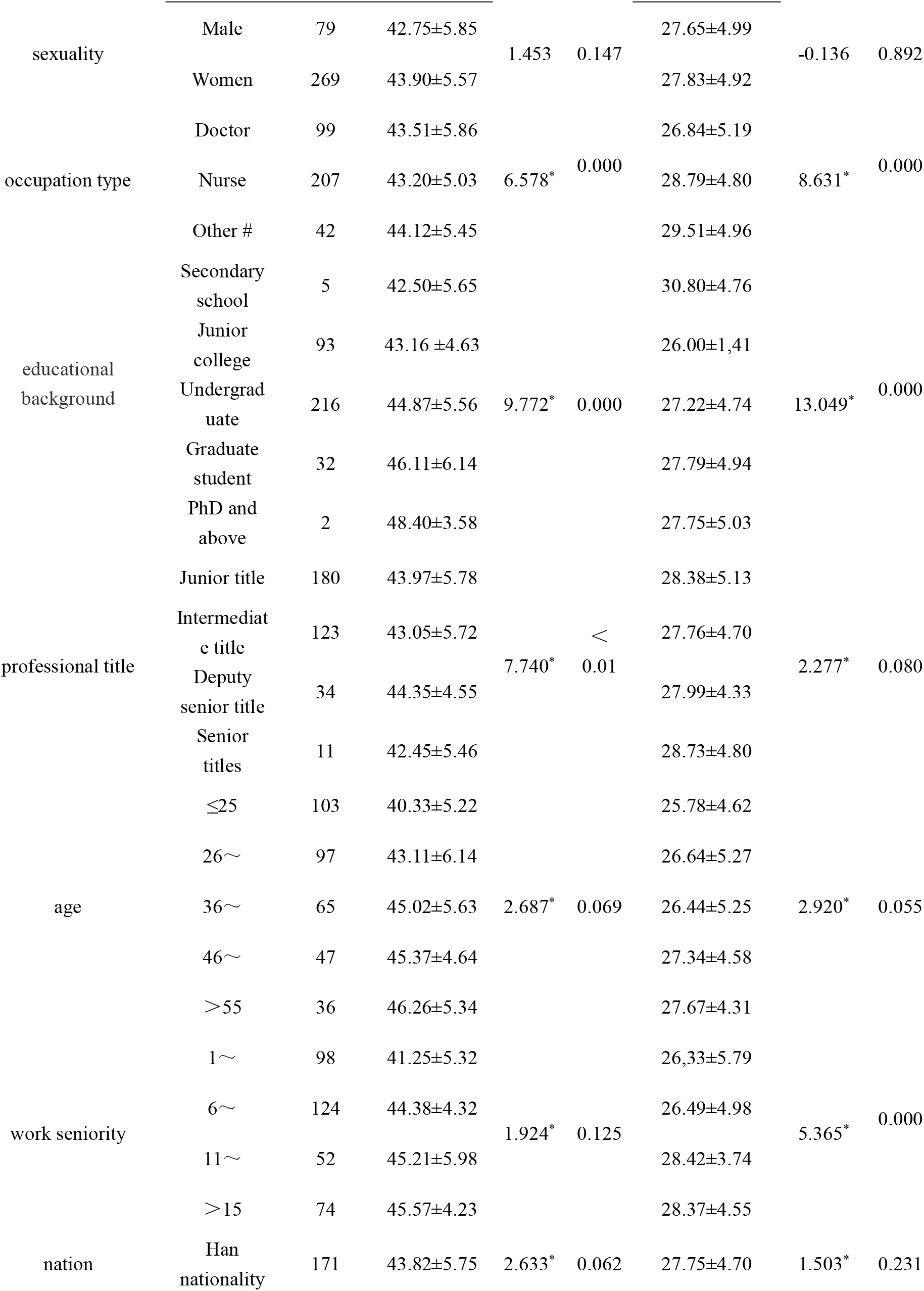
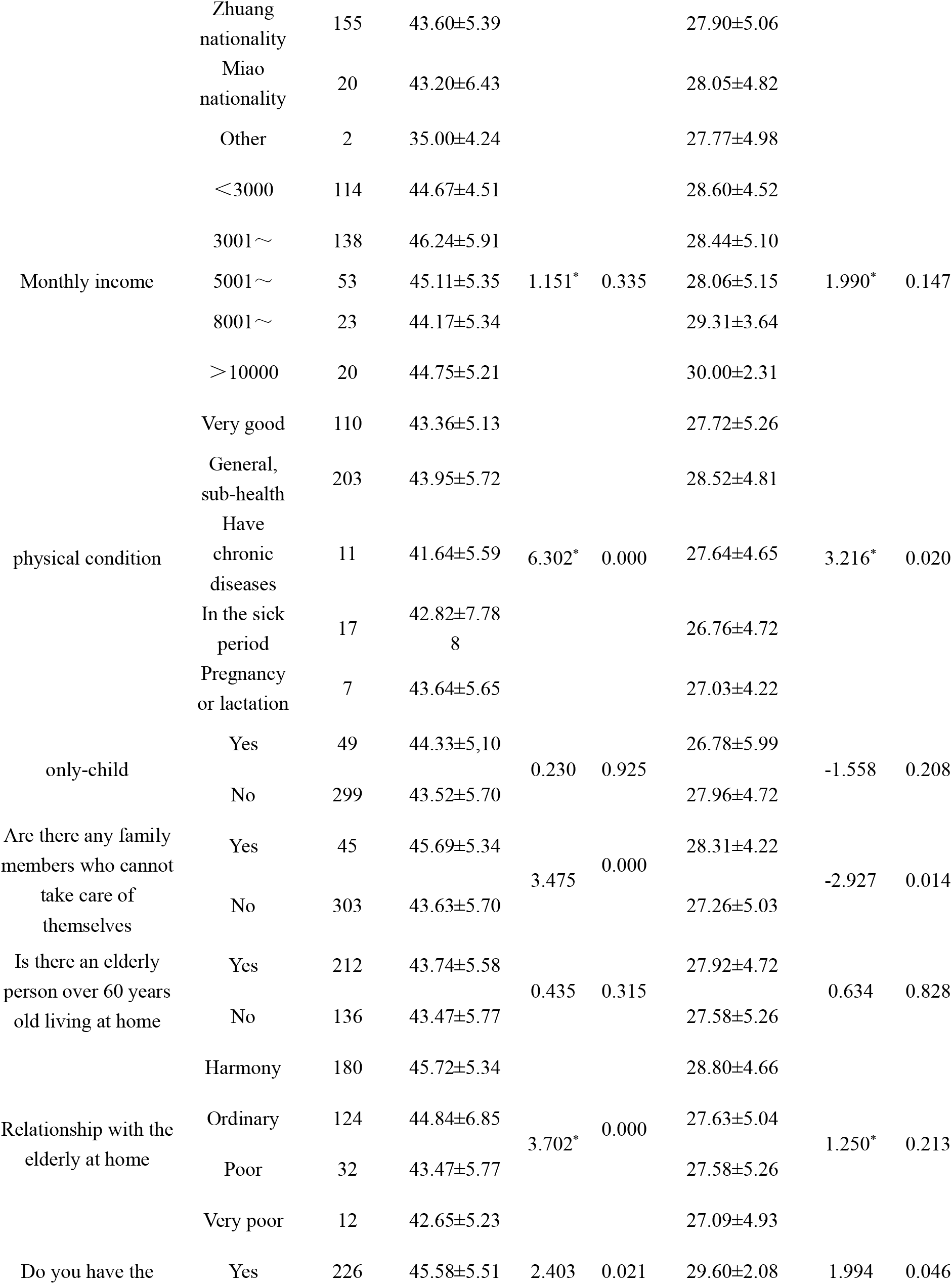
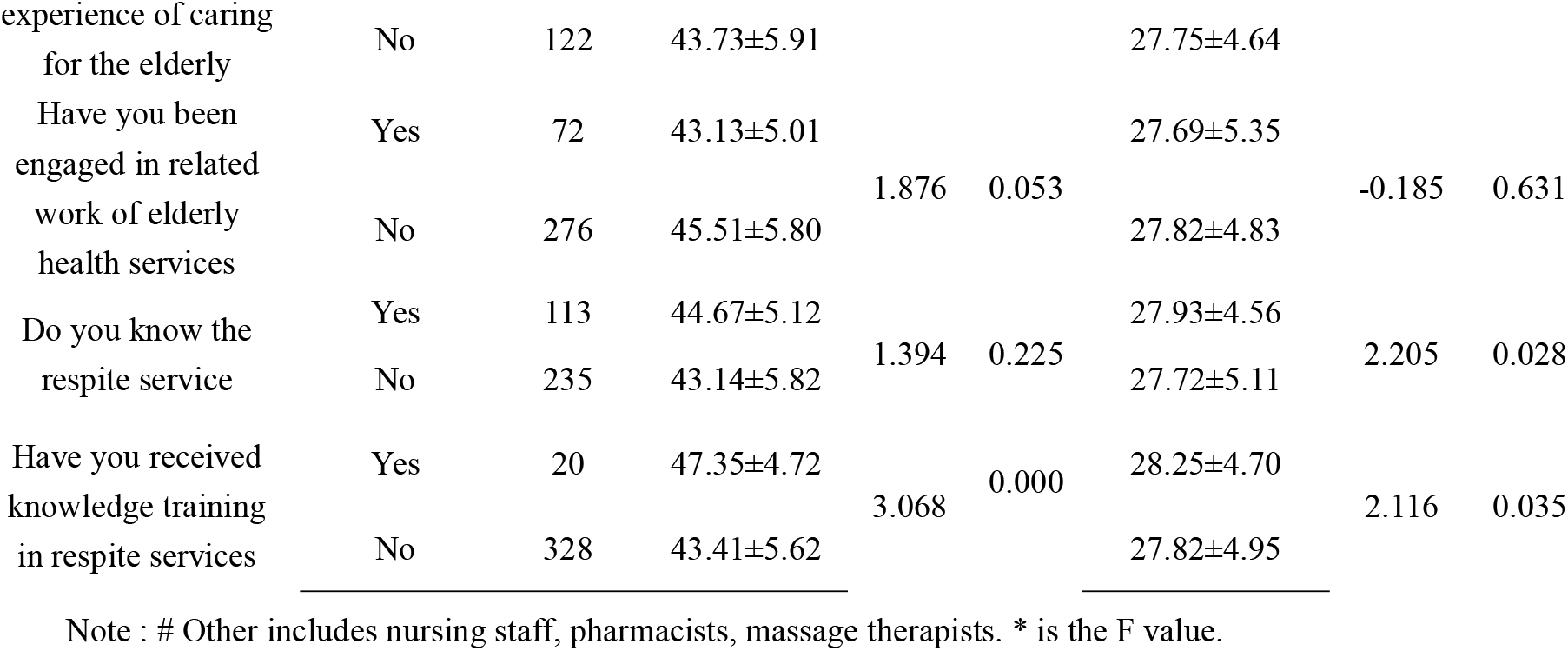
Single factor analysis of influencing community medical staff’s respite services attitude and behavioral intention score.

### 3.3 Multiple linear regression analysis of the influence of community medical staff attitude and behavior intention score

The scores of medical staff’s attitude and behavior intention were taken as dependent variables, and the factors with statistical significance in single factor analysis were taken as independent variables for multiple stepwise regression analysis. The variable assignment is shown in Table 3. The results of multivariate linear regression analysis showed that occupation, education, whether there were family members who could not take care of themselves due to major diseases, whether there was experience in caring for the elderly, the relationship with the elderly at home, and whether they had received knowledge training in respite service were the influencing factors of community medical staff’s attitude towards respite service ( P < 0.05 ). Education background, working years, whether there are family members who cannot take care of themselves due to major diseases, whether there is experience in caring for the elderly, whether they know about respite service, and whether they have received knowledge training on respite service are the influencing factors of behavioral intention of community medical staff on respite service ( P < 0.05 ). Table 4 and Table 5.

**Table 3.** Variable assignment table.

| factor | Description of valuation |
| --- | --- |
| Profession | Doctors = 1, nurses = 2, others = 3 |
| Education | Secondary school = 1, junior college = 2, undergraduate = 3, postgraduate = 4, doctoral and above = 5 |
| Professional title | Primary = 1, intermediate = 2, secondary = 3, advanced = 4 |
| Years of work | Measured value |
| Your own physical condition | Very good = 1, general = 2, sub-health = 3, chronic disease = 4, in sick = 5, pregnancy or lactation = 6 |
| Are there family members with major diseases who can not take care of themselves | Yes = 1, no = 2 |
| Relationship with the elderly at home | Harmony = 1, Ordinary = 2, Poor = 3, Very Poor = 4 |
| Do you have the experience of taking care of the elderly | Yes = 1, No = 2 |
| Do you know the respite services | Yes = 1, No = 2 |
| Have you received knowledge training in respite services | Yes = 1, No = 2 |

**Table 4.** Multiple linear regression analysis of influencing community medical staff’s attitude towards respite services.

| factor | $\beta$ | SE | Wald B | t | P |
| --- | --- | --- | --- | --- | --- |
| Constants | 13.615 | 1.627 | — | 8.481 | 0.000 |
| Profession | 0.621 | 0.186 | 0.189 | 3.176 | 0.002 |
| Education | 0.418 | 0.121 | 0.136 | 3.721 | 0.000 |
| Title | 0.165 | 0.287 | 0.015 | 0.574 | 0.566 |
| Are there family members with major diseases who can not take care of themselves | -0.231 | 0.057 | -0.066 | -3.365 | 0.001 |
| Your own physical condition | -0.442 | 0.195 | -0.052 | -1.268 | 0.223 |
| Relationship with the elderly at home | -0.275 | 0.085 | -0.163 | 3.150 | 0.002 |
| Do you have the experience of taking care of the elderly | 0.287 | 0.089 | -0.123 | 3.462 | 0.000 |
| Have you received knowledge training in respite services | -0.868 | 0.092 | -0.201 | -6.637 | 0.000 |

Note : $R^2 = 0.571$ , adjusted $R^2 = 0.553$ , $F = 31.211$ , $P = 0.000$ .

**Table 5.** Multiple linear regression analysis of the influence of community medical staff on the behavior intention of respite services.

| factor | $\beta$ | SE | Wald B | t 值 | P 值 |
| --- | --- | --- | --- | --- | --- |
| Constants | 28.377 | 0.369 | - | 56.752 | 0.000 |
| Profession | 0.231 | 0.203 | 0.010 | 0.967 | 0.538 |
| Education | 1.363 | 0.431 | 0.316 | 3.125 | 0.000 |
| Years of work | 3.421 | 1.146 | 0.182 | 3.027 | 0.002 |
| Are there family members with major diseases who can not take care of themselves | -1.101 | 0.483 | -0.118 | -2.301 | 0.023 |
| Your own physical condition | -0.009 | 0.152 | -0.001 | -0.059 | 0.953 |
| Do you have the experience of taking care of the elderly | -0.188 | 0.069 | -0.053 | -2.485 | 0.018 |
| Do you know the respite services | -0.856 | 0.076 | -0.245 | -7.458 | 0.000 |
| Have you received knowledge training in respite services | 0.388 | 0.172 | 0.100 | 2.197 | 0.030 |

Note : $R^2 = 0.427$ , adjusted $R^2 = 0.315$ , $F = 10.231$ , $P = 0.000$ .

## 4. Discussion

The development of respite services in China started relatively late, and it is still in the initial stage of pilot. The results of our study show that the attitude and behavioral intention scores of community medical staff on respite services are low, similar to the research of Castro AR[16]. The attitude items with lower scores are mainly 68.3 % of community medical staff think that ‘the income and pay of respite service are not proportional’, 61.5 % think that ‘providing home respite service is unsafe for our medical staff’ and 52.8 % of medical staff think that ‘providing respite service makes me feel that my own value is not reflected’. The reason may be that the current respite service as a new way of old-age service, the service policy, service mode, service system and service standard of respite service are in the exploratory stage, and the staff assessment and incentive mechanism for providing respite service are not perfect, which dampens the enthusiasm of the staff[17]. Studies have shown that one of the reasons that affect the use of respite services is because staff do not understand the way, meaning and role of respite services[18, 19]. In our study, only 64.66 % of the community medical staff know the respite services, and only 5.75 % of the people have received the training of the respite services. Therefore, the relevant departments need to carry out multi-level publicity of the respite services and improve the training of professional talents, especially medical staff, nursing staff of pension institutions, social workers, etc., so that they can have a deep understanding and understanding of the respite services, accept the respite services from the ideological concept, so as to change the attitude towards the respite services, and then improve the action force. At the same time, we should enhance the professional identity of respite service personnel from the perspectives of improving salary, establishing incentive mechanism and multi-channel publicity[20].

In the analysis of the influencing factors of attitude and behavior intention, we found that the factors that have a significant impact are concentrated in three dimensions : knowledge structure, personal experience and professional training. The first is the knowledge structure and cognitive level. As a manifestation of personal comprehensive cognitive ability, education has a positive predictive effect on attitude and behavioral intention. Highly educated people usually have stronger information integration and critical thinking ability, can more deeply understand the long-term value of respite services in public health and health management[21, 22], and are more likely to actively learn and study new knowledge. Therefore, the development and promotion of respite services need to pay attention to the driving role of highly educated and highly qualified staff. It is worth noting that ‘whether you know respite services’ is a unique influencing factor of behavioral intention. This further shows that awareness is the logical starting point of behavioral intention[23]. The second is the deep empathy of personal life experience. The presence of a person in need of care at home is the most powerful driving force for the intrinsic identification of medical personnel. Whether they have a family member who is unable to take care of themselves due to a major disease at home, or have their own experience of caring for the elderly, this ‘dual role’, both as a professional provider and as a family caregiver or potential caregiver, enables them to experience the enormous physical and mental stress and financial burden of long-term care[24]. Therefore, empathy is a key tool to promote understanding and trust between doctors and patients, and an important guarantee for a harmonious doctor-patient relationship. Managers should pay attention to the training of medical staff’s empathy ability and improve their empathy ability[25, 26]. The third is the key empowerment of systematic professional training. Having received knowledge training is the most critical bridge connecting attitude and behavioral intention. Systematic knowledge training, such as service process, communication skills, risk control, resource links, etc., can improve the self-efficacy of medical staff. Training will consolidate and enhance positive attitudes by eliminating uncertainty, and more directly and effectively drive behavioral intentions[27].

At the same time, we found that the influencing factors of attitude are more focused on emotion and subjective feelings, and the type of occupation and the relationship with the elderly at home significantly affect attitude rather than behavioral intention. This may be due to the fact that the harmonious family relationship itself shapes its positive emotional tone towards elderly care, which is transferred to the subjective will of all elderly support services[28, 29]. Different types of occupations have different value judgments on services due to different work focuses, but this difference may not be directly converted into a difference in willingness to act[30]. The influencing factors of behavioral intention emphasize more on experience and practical feasibility, and working years are the unique influencing factors of behavioral intention. Senior medical staff have richer clinical experience and a broader social perspective, and may know more clearly how to mobilize resources and avoid risks to promote one thing. The practical confidence brought by this experience directly strengthens their behavioral intention[31]. Most importantly, the high consistency of the two influencing factors strongly confirms the applicability of the theoretical framework of knowledge-attitude-practice in the context of our study. Positive attitude is a necessary prerequisite for positive behavioral intentions, and systematic knowledge training and personal deep empathy experience are the key catalysts to promote the transformation from attitude to behavior.

## 5. Conclusions

The attitude of community medical staff to respite service is not positive, and the behavioral intention is low. It is necessary to strengthen the training of respite services knowledge for medical staff with low educational background and working years, strengthen the cultivation of empathy and empathy ability of medical staff, carry out multi-level publicity of respite services, and improve people’s awareness and acceptance of respite services, so as to promote the development of respite services industry and promote healthy aging. Our study only conducted a sample survey in Guangxi, and the sample size is relatively small. In the future, a large-scale and large-sample study is needed to further explore the attitudes and behaviors of community medical staff on respite services.

## Data Availability

The datasets used and analyzed in the present study are available from the corresponding author on reasonable request.

## Notes

### Competing Interest Statement

The authors have declared no competing interest.

### Funding Statement

Guangxi Medical University Youth Science Fund Project GXMUYSF202379

### Author Declarations

The study was reviewed and approved by the Medical Ethics Committee of The Second Affiliated Hospital of Guangxi Medical University (No. 166 Daxue East Road, Nanning 530007, Guangxi, China) under approval number 2023-KY-0921.

